# Role of Lifestyle and Risk Factor Modification Clinics in Patients with Atrial Fibrillation: A Systematic Review and Meta-Analysis of Randomised Controlled Trials

**DOI:** 10.64898/2026.03.16.26348558

**Authors:** Yan Zhao, Francis J Ha, Adam J Brown, Nitesh Nerlekar

## Abstract

**Background:** Incidence and recurrence of atrial fibrillation (AF) is associated with several lifestyle risk factors. Lifestyle and risk factor modification (LRFM) clinics could have a role in comprehensively addressing AF from a holistic patient-centred approach to improve clinical outcomes.

**Methods:** We performed a systematic review and meta-analysis of randomised controlled trials (RCTs) evaluating the role of LRFM clinics compared with usual care (UC) in patients with AF. The primary endpoint was atrial arrhythmia recurrence. Secondary endpoints were AF and heart failure (HF) related hospitalisation, cardiovascular death, stroke or transient ischaemic attack (TIA), and quality-of-life (QOL).

**Results:** A total of eleven RCTs with a total of 3364 patients were included (five RCTs performed in the context of AF ablation). Mean age was 58–73 years, 30% were female and 18% had persistent AF. Duration of follow-up ranged from 3-24 months. LRFM clinics significantly reduced the primary endpoint of arrhythmia recurrence compared with UC after catheter ablation (OR 0.34, 95% CI 0.23-0.51, p<0.001, I^2^=0%). LRFM clinics also reduced AF-related hospitalisation (OR 0.70, 95%CI 0.51–0.98, p=0.04, I^2^=21%) and improved QOL (mean improvement on Short Form 36 Questionnaire 8.90, 95% CI 7.6.91–10.90, p<0.001). There was no difference between LRFM clinics and UC for HF-related hospitalisation (p=0.16), cardiovascular deaths (p=0.79) or stroke/TIA (p=0.83).

**Conclusion:** In this meta-analysis of RCTs, LRFM clinics reduced AF recurrence after ablation, reduced AF-related hospitalisation and improved QOL. This study supports a comprehensive multidisciplinary lifestyle risk modification model of care to improve clinical outcomes in patients with AF.

## INTRODUCTION

The burden of atrial fibrillation (AF) is growing worldwide.(1) Beyond a disease state itself, AF is a prognostic marker for increased cardiovascular risk including stroke and heart failure (HF).(2) Lifestyle factors such as weight loss, physical exercise and alcohol abstinence are well recognised to influence the incidence and recurrence of AF. (3–5) It is likely these factors would cumulatively impact clinical outcomes related to AF when addressed. As such, there is an unmet need to understand the role of comprehensive lifestyle and risk factor modification clinics (LRFM) that address these upstream factors related to AF, patient education and medication adherence. This has been predominantly studied in the context of nurse-led clinics, sometimes coupled with a multidisciplinary team providing tailored lifestyle interventions in a patient-focused approach.(6,7) Recent randomised controlled trials (RCTs) have evaluated this approach in the context of AF ablation.(8–10) Beyond a holistic approach to patient health beyond AF, these comprehensive multidisciplinary clinics could also reduce the onus on clinical electrophysiologists alone in being able to adequately address certain lifestyle behaviours within the practical time constraints of outpatient care.

We thus performed a systematic review and meta-analysis of randomised controlled trials (RCTs) to evaluate the role of LRFM clinics in the management of AF in general as well as in the context of AF ablation.

## METHODS

Online searches on major electronic databases for RCTs were conducted including PubMed, Scopus, Embase and ClinicalTrials.gov, using a combination of the following search phrases: (“nurse led” OR “nurse clinic” OR “nurse intervention” OR “nurse care”) AND (“atrial fibrillation” OR “AF”). Given the broad spectrum of lifestyle and risk factor modification in the context of AF, we added a specific search string of (“Randomised trials”) AND (“lifestyle management clinic” OR “lifestyle modification clinic”) AND (“atrial fibrillation” OR “AF”). An additional manual search on presentations and abstracts was conducted on recent major international conferences. Studies up to 6^th^ Nov 2025 were included in the searches. The study protocol was registered with PROSPERO (CRD420251163105).

Inclusion criteria for the review were: 1) RCTs that investigated LRFM clinics compared with usual care (UC) in patients with AF; 2) LRFM clinics that encompassed comprehensive risk factor and lifestyle assessment; 3) nurse-or physician-led multidisciplinary clinics in the treatment group; 4) studies in humans; and 5) studies published in English. Exclusion criteria were studies that only evaluated single risk factor (e.g. weight loss alone, alcohol abstinence alone), and studies involving non-randomised comparisons.

### Endpoints

The primary endpoint was recurrence of any atrial arrhythmia recurrence at follow-up. Secondary endpoints were AF-related hospitalisation, heart failure (HF) related hospitalisation, their pooled hospitalisation (AF and HF), cardiovascular (CV) death, stroke or transient ischaemic attacks (TIA), and qualify-of-life (QOL).

### Screening and Data Extraction

Two reviewers carried out screening and data extraction process independently. Firstly, study description items were extracted, including location and duration of the study, number of participants, type of intervention and follow-up duration. Study characteristics were collected including patient baseline characteristics and cardiovascular co-morbidities. Clinical outcomes with number of events in each assigned group, and QOL measures such as Short Form 36 Questionnaire (SF36) scores were also extracted.

### Quality Assessment

Risk of bias was assessed against the revised Cochrane risk of bias tool for randomised trials (RoB 2), 2019 version (11). Summary of risk of bias assessment is provided **Table S1** in the Data Supplement.

### Statistical Analysis

Data analyses were conducted on Cochrane RevMan. Random-effect modelling was used for all subgroups. Statistics are reported as pooled odds ratios (OR) with 95% confidence intervals (CI) for dichotomous datasets such as number of events, and mean difference with 95% confidence intervals (CI) for continuous variables. Funnel plots were utilised to visually assess for publication bias.

## RESULTS

### Search Results

Initial search on various databases yielded a total number of 663 studies. After duplicate removal, two independent reviewers carried out initial screening of 414 studies by title and abstract for eligibility (**Figure 1**). After exclusion of studies found to be irrelevant or outside of the scope of this review (401 studies), a total of 11 RCTs were included in the final systematic review and meta-analysis. (6–10,12–17)

**Figure 1.**
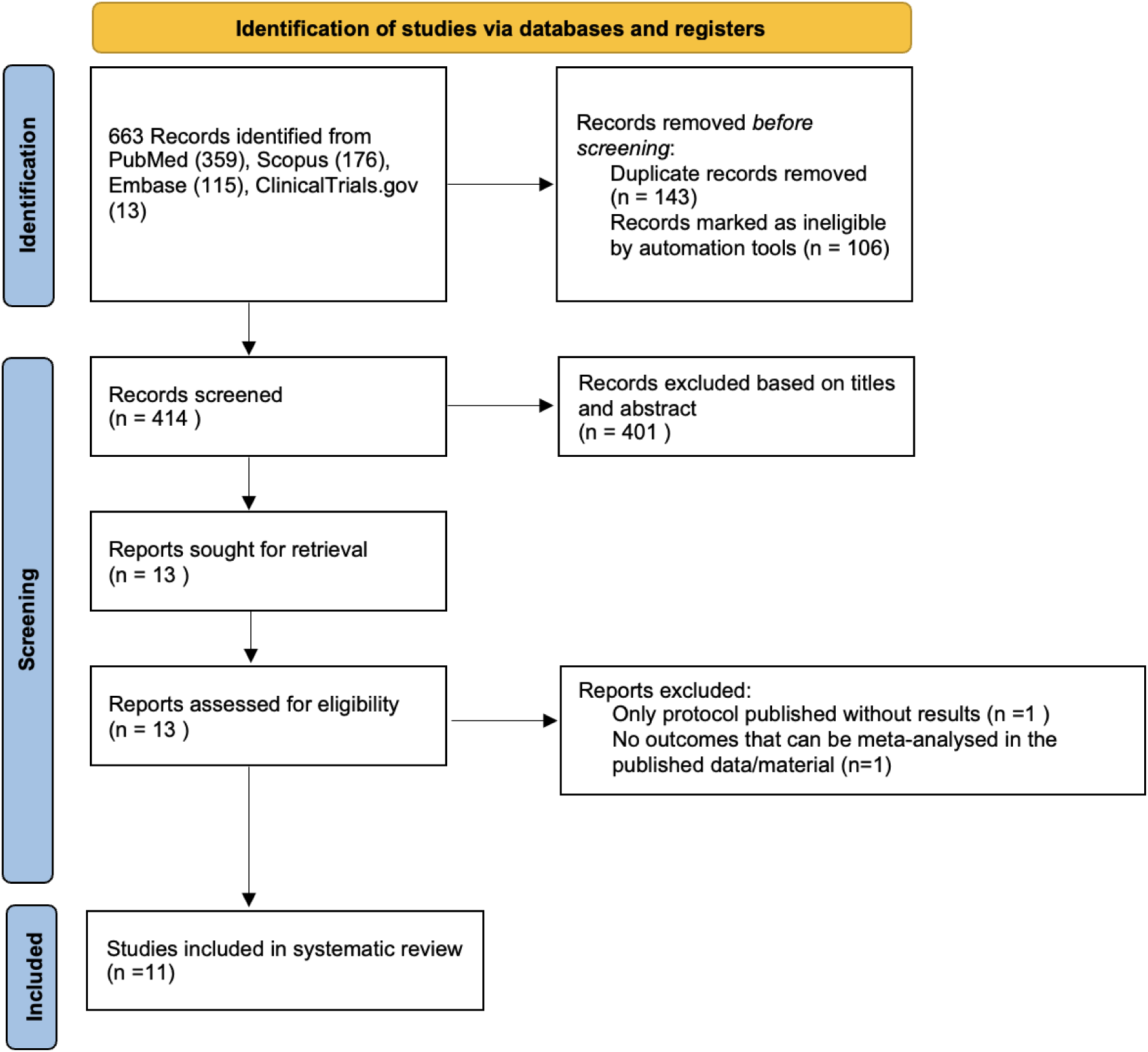
PRISMA diagram.

### Characteristics of trials

Across 11 RCTs there were a total of 3364 patients with 1682 (50%) in LRFM group and 1682 (50%) in the UC group. Mean age ranged from 58-73 years, 30% (1009/3364) were female and 18% (610/3364) had persistent AF. Duration of follow-up ranged from 3 to 24 months. Five RCTs were conducted in the context of AF ablation (558 patients, 17%). Ten RCTs (6–10,12,14–17) involved nurse-led LRFM clinics as their intervention group, and one (13) was physician-led. Medication profile of participants, and detailed LRFM clinic structure of each trial are shown in Table S2 and Table S3 in the supplementary material.

### Primary Endpoint

Five RCTs reported atrial arrhythmia recurrence as their primary end after AF ablation.(8–10,13,17) LRFM clinics significantly reduced risk of atrial tachyarrhythmia recurrence compared with UC after AF ablation (OR 0.34, 95% CI 0.23 – 0.51, p<0.001, I^2^=0%; **Figure 2**).

**Figure 2.**
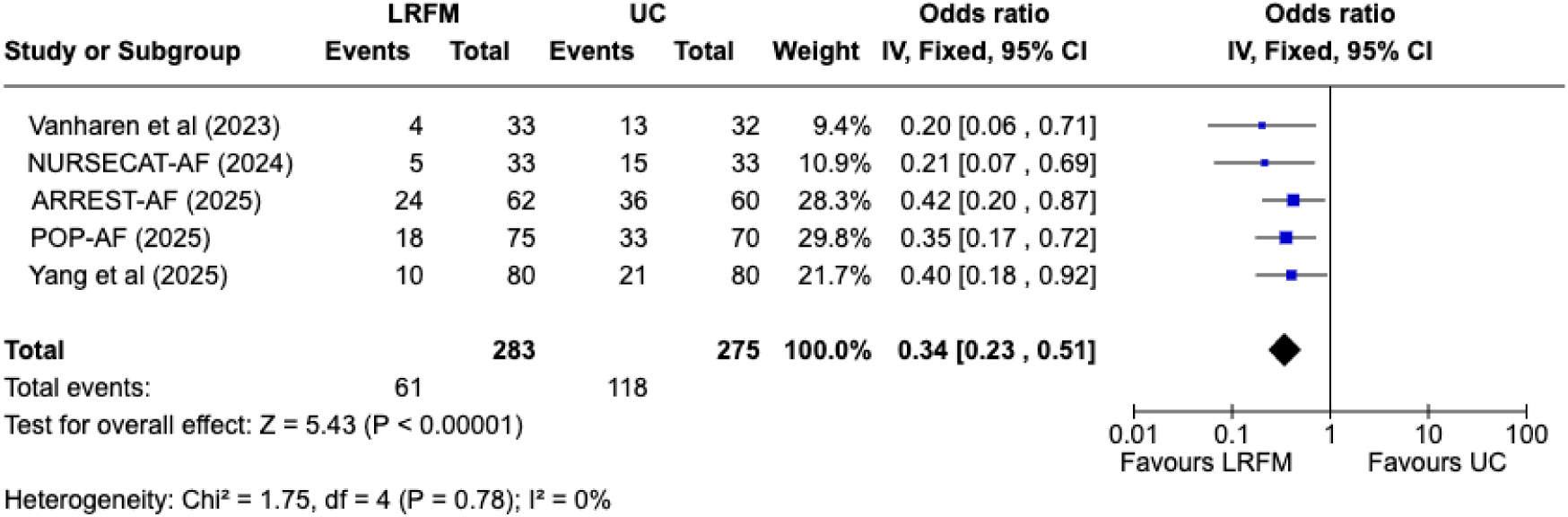
Forest plot of atrial arrhythmia recurrence after catheter ablation comparing lifestyle risk factor modification clinic with usual care.

### Secondary Endpoints

#### Hospitalisation

Five trials reported hospitalisation as an endpoint in a general AF population, while one trial (POP-AF trial) reported HF hospitalisation after AF ablation. (6,8,12,14,16) LRFM clinics significantly reduced pooled hospitalisation (AF and HF) compared with UC in the general AF population (OR 0.74, 95% CI 0.59-0.91, p=0.004, I^2^=0%; **Figure 3**). Furthermore, LRFM clinics reduced AF-related hospitalisation compared with UC in the general AF population (OR 0.70, 95% CI 0.51- 0.98, p=0.04, I^2^=21%) although did not significantly reduce HF hospitalisation in the general AF population (OR 0.71, 95% CI 0.44-1.15, p=0.16).

**Figure 3.**
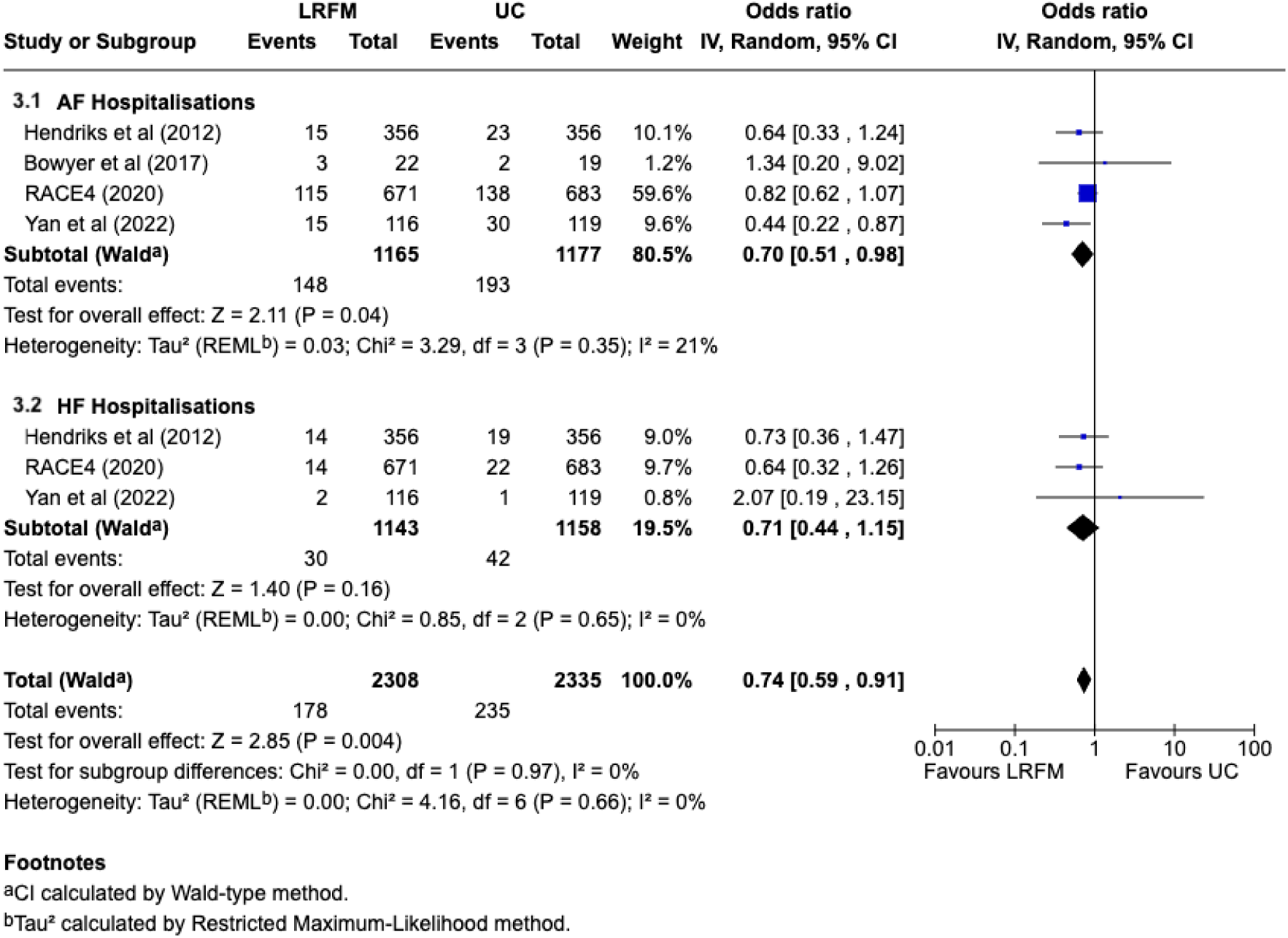
Forest plot of atrial fibrillation and heart failure hospitalisation in general AF population comparing lifestyle risk factor modification clinic with usual care.

#### Cardiovascular death and stroke/transient ischaemic attack

Four trials assessed CV deaths and/or stroke/TIA with one trial reporting (8) post AF ablation.(6,8,14,16) LRFM clinics did not reduce risk of CV death (general AF: OR 0.81, 95% CI 0.18-3.71, p=0.79, I^2^=63%; post AF ablation: OR 0.31, 95% CI 0.01 – 7.66, p = 0.47), or stroke/TIA (general AF: OR 1.08, 95% CI 0.56-2.08, p=0.83, I^2^=0%; post AF ablation: OR 0.18, 95% CI 0.01 – 3.85, p = 0.27) compared with UC (**Figure 4**).

**Figure 4.**
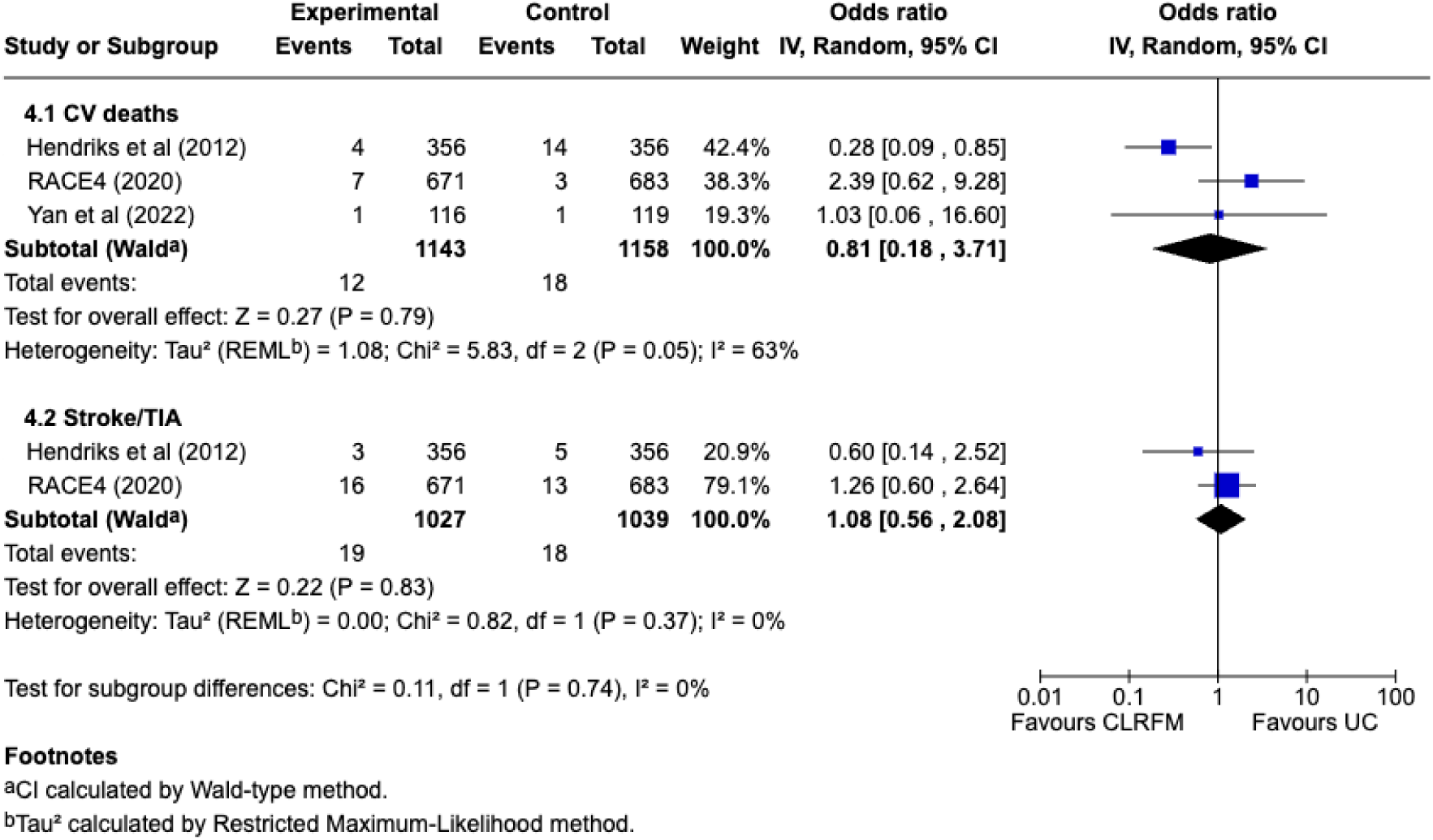
Forest plots of cardiovascular deaths and stroke/transient ischaemic attacks in general AF population comparing lifestyle risk factor modification clinic with usual care.

#### Quality of life

Seven trials (7,10,12–15,17) evaluated QOL outcomes with three trials using SF36 score (10,12,14), and the other trials using Atrial Fibrillation Effect on Quality-of-Life Questionnaire (AFEQT) score (7), Arrhythmia-Specific Scale in Tachycardia and Arrhythmia (ASTA) (17), and Atrial Fibrillation Severity Scale (AFSS) (13). A meta-analysis was performed of RCTs that used the SF36 score (**Figure 5**). (10,12,14) LRFM clinics significantly improved overall SF36 score with a mean difference of 8.80 (95% CI 7.90–9.69, p<0.001). Individual components were improved including physical functioning (mean difference 9.49, 95% CI 7.58–11.40, p<0.001), social functioning (mean difference 6.25 95% CI 4.32–8.18, p<0.001), general health (mean difference 8.40, 95% CI 6.41–10.39, p<0.001), vitality (mean difference 8.62, 95% CI 6.66-10.58, p<0.001), physical role limitation (mean difference 15.86, 95% CI 8.73–22.98, p<0.001), emotional role limitation (mean difference 8.42, 95% CI 1.35–15.49, p = 0.02), mental health (mean difference 13.52, 95% CI 10.21–16.84, p < 0.001) and bodily pain (mean difference 10.24, 95% CI 5.89–14.59, p<0.001). Additionally, three RCTs showed improvement in QOL with LRFM clinics as measured by AFEQT score (7), ASTA (17), and AFSS (13).

**Figure 5.**
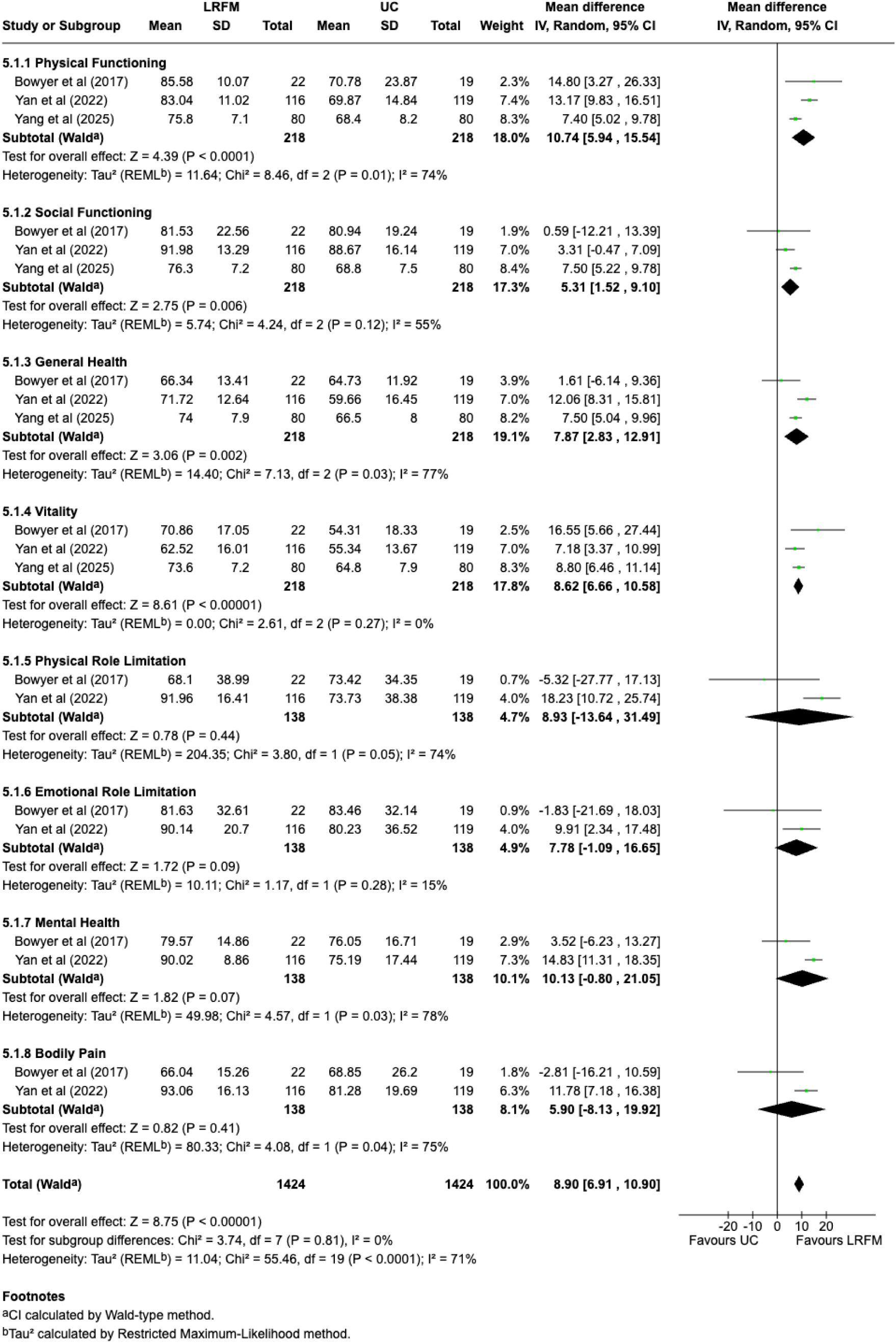
Forest plot of components of Short Form 36 Questionnaire (SF 36) comparing lifestyle risk factor modification clinic with usual care.

#### Publication Bias

There was no publication bias observed in the trials included for any primary or secondary endpoints on visual examination of funnel plots as shown in **Figure 6**.

**Figure 6.**
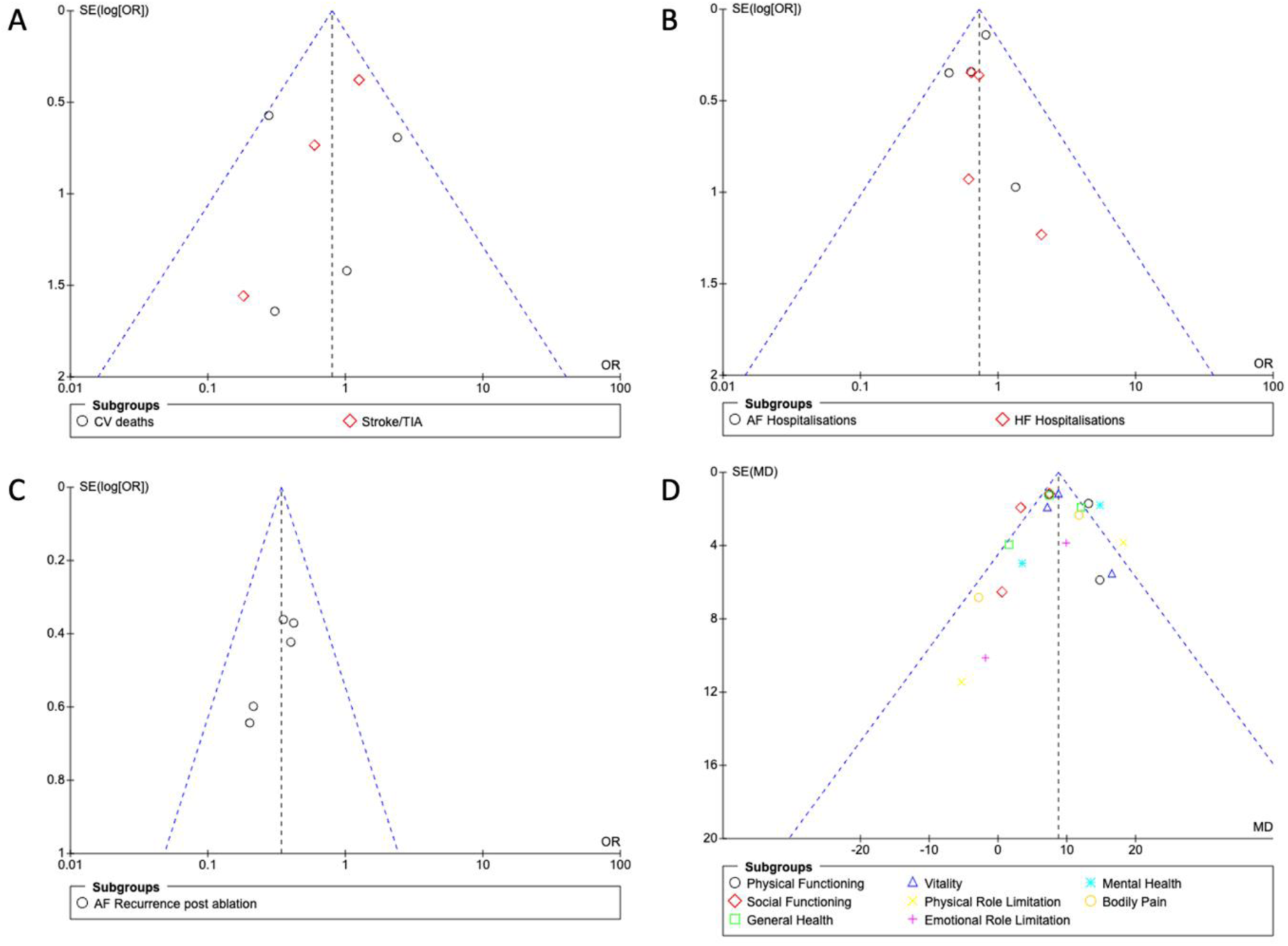
Funnel plot for various outcomes examined in this analysis. (A) Cardiovascular death and stroke/transient ischaemic attack (B) atrial fibrillation and heart failure hospitalisation (C) Atrial tachyarrhythmia recurrence post ablation (D) SF36 components at last follow-up.

## DISCUSSION

We performed a systematic review and meta-analysis of RCTs to determine the role of LRFM clinics in the management of AF. We found that that LRFM clinics reduced atrial tachyarrhythmia recurrence after ablation, reduced AF-related hospitalisation and improved QOL. There was no significant difference with regards to HF hospitalisation, CV deaths or stroke/TIA compared to UC.

Randomised controlled trials consistently showed a significant reduction in AF recurrence after ablation when assigned to LRFM clinics over a follow-up period ranging from 6 to 12 months.

These data are crucial in view of certain limitations of catheter ablation alone in reducing arrhythmia recurrence. While the advent of pulsed field ablation (PFA) appears promising in terms of procedural workflow with no evidence of oesophageal injury thus far,(18,19) arrhythmia outcomes have still been similar to thermal ablation thus far.(20) This highlights the inherent deficit of addressing AF from a unidimensional model of medical treatment alone and there is an ongoing need to address upstream risk factors. Of note, these trials also highlight the opportunity to impact clinical outcomes during a relatively short duration of follow-up. Patients should be counselled with regards to augmented effects of AF ablation when coupled with lifestyle modification. Additionally, this shifts the focus from AF itself to a holistic approach that has been associated with greater patient satisfaction, knowledge of their AF and medication adherence.(7,9) This model of integrated care is pertinent given the substantial benefits observed not only for AF related outcomes but also their general physical health including weight reduction and lower systolic blood pressure.(13)

Health care expenditure related to AF is rapidly growing. In the United States, patients with AF have a roughly 1.6 times greater annual total health care cost compared with those without AF, mostly related to inpatient visits and prescriptions.(21,22) Expenditure is also currently acutely increased in view of the efficiency and workflow of PFA that has high equipment cost at present compared with conventional thermal ablation.(23) Current systems of healthcare that revolve around inpatient management, procedures (e.g. AF ablation) and prescriptions do not address AF holistically. This includes the role of upstream lifestyle behaviours that significantly influence clinical outcomes. Indeed, the POP-AF trial showed significantly fewer repeat ablations and direct current cardioversions in patients randomised to nurse-led LRFM clinic in combination with their AF ablation.(8) This was a profoundly holistic approach that included a multidisciplinary assessment of lifestyle behaviours in a structured interview (physical inactivity, alcohol consumption, smoking), hypertension, lipid and glycaemic management and obstructive sleep apnoea assessment. While service provision is contingent on each country’s health care system and individual centre capacity, such a model of care is evidence that integrated LRFM clinics for AF can have demonstrable benefits in reducing hospitalisation.

One of the challenges in delivering LRFM clinics across these trials is the heterogeneity and variation in the actual model of care. In general, they involve a structured interview and educational session that addresses lifestyle factors, education about AF and the role of medications and adherence. Cardiovascular screening including lipids, glycaemic control, blood pressure and obstructive sleep apnoea may also be integrated.(8) Certain trials involved a multidisciplinary team comprising cardiologists, pulmonologists, sleep medicine specialists, endocrinologists, their general practitioner, dietician, physiotherapist, psychologist and smoking cessation counsellor.(8,10) The time commitment both from a staff and patient perspective is also variable with a mixture of telephone calls, in-person follow-up visits every 3 months. The LRFM clinic intervention can last up to 6 months prior to offering an ablation.(8,12,16) Dedicated software guiding comprehensive management of AF and other associated cardiovascular conditions based on individual patient symptoms, type of AF and stroke risk has sometimes been integrated.(16) Practicalities of service delivery must be carefully considered by each individual health service.

The role of LRFM clinics in influencing hard endpoint such as CV death or stroke in the context of AF has been conflicting. A large trial from the Netherlands reported reduced CV death (1.1% versus 3.9%; hazard ratio 0.28) in patients randomised to nurse-led LRFM clinics for outpatient management of AF compared with UC.(16) In a post-hoc analysis of the same trial at longer follow-up (22 months), integrated nurse-led LRFM clinics was associated with reduced all-cause death although this was primarily driven by reduced CV death.(24) Conversely, another larger trial of 1375 patients from the Netherlands found a numerically higher risk of CV death in patients randomised to LRFM care.(6) Practically, It would be challenging to detect differences in hard end points at short term follow-up from LRFM clinics. The CABANA trial failed to show any difference in all-cause death or stroke in patients randomised to AF ablation compared with medical therapy in 2204 patients after a median follow-up of over four years.(25) It is conceivable that in a higher risk population, such as those with HF with reduced ejection fraction, the implementation of LRFM clinics could influence mortality. However, this remains uncertain at present and large prospective studies with long term follow-up are needed to assess this further.(26)

We acknowledge several limitations in this study. First, there is heterogeneity in implementation and duration of LRFM clinics across trials. How these data are extrapolated to various healthcare systems is not yet clear. Arguably the consistent results from a meta-analysis provides evidence for broader applicability than an individual study in isolation. Second, endpoints varied between trials, especially pertaining to certain QOL and medication adherence measures that limits direct comparability between certain trials. Third, patients enrolled in these clinical trials reflect a certain population who are potentially self-selected towards being more motivated and agreeable to undertaking lifestyle modifications.

## CONCLUSION

In this systematic review and meta-analysis, LRFM clinics reduced atrial arrhythmia recurrence after ablation and hospitalisation and improved QOL. This study supports a multidisciplinary comprehensive lifestyle and risk modification model of care to improve clinical outcomes and address certain resource limitations with the increasing burden of AF.

## Data Availability

This is a systematic review and meta-analysis. All data were from published studies.

## Acknowledgement

None of the authors have any conflict of interest to disclose, nor have received any funding or grant from any organisations. No artificial intelligence was used in this systematic review and meta-analysis.

---

**Table 1.** Characteristics of randomised controlled trials.

| Trial (publication year) | Location | Duration | No. of participants | Follow up | Lifestyle and risk factor modification clinic description | AF/AFL/AT recurrence | Hospitalisation | CV death and stroke/TIA | QOL |
| --- | --- | --- | --- | --- | --- | --- | --- | --- | --- |
| Hendriks et al. (2012) | Netherlands | 2007 - 2008 | 712 | 3,6,12 months, then 6 monthly | 30 min outpatient nurse-led consultation that provides education and management guided by software CardioConsult AF | No | Yes | Yes | No |
| Bowyer et al. (2017) | Australia | NR | 41 | 6 months | Nurse-led education sessions on AF causes, risk factors, goals of treatment and lifestyle modification | No | Yes | No | Yes (SF36) |
| RACE4 (2020) | Netherlands | 2012-2017 | 1375 | 3,6,12 months, then yearly | Nurse-led outpatient consultation by using guidelines-based decision-support software, ensuring management of AF is followed + provision of psychosocial support + education | No | Yes | Yes | No |
| NEAT (2020) | Australia | 2015-2017 | 72 | 3 months | Nurse-led educational and risk factor management session including medication adherence and cardiovascular risk factor profiling, health counselling and goal setting. | No | No | No | Yes (SF12) |
| Yan et al (2022) | China | 2018-2020 | 235 | 1,3,6,12 months, PRN thereafter | Nurse-led interviews include educational and psychological support, care coordination and liaison | No | Yes | Yes | Yes (SF36) |
| Vanharen et al, (2023) | Belgium | 2021 - NR | 65 | 6 months | Nurse-led education on physical activity, alcohol intake, weight reduction, smoking cessation, adherence to DOACs | Yes | No | No | No |
| NURSECAT-AF (2024) | Spain | 2022-2024 | 66 | 12 months | Nurse-led educational programme on AF, support for patient undergoing ablation, risk factors management | Yes | Yes (readmission of no specified cause) | No | Yes (ASTA) |
| ARREST-AF (2025) | Australia | 2014-2017 | 122 | 3 monthly for 12 months | Physician-led comprehensive lifestyle and risk factor management clinic for BP, weight, lipid management, glycaemic control, sleep-disordered breathing management, smoking and alcohol consumption | Yes | No | No | Yes (AFSS) |
| Li et al. (2025) | Hong Kong | 2019-2022 | 392 | Immediately after intervention, and 6 months | Nurse-led risk profile assessment, empowered shared decision making of DOAC use, empowered AF self-management and increased access to professional advice. | No | No | No | Yes (AFEQT) |
| POP-AF (2025) | Netherlands | 2021-2024 | 162 | 3 and 12 months | Nurse-led Extensive risk factor assessment and lifestyle modification intervention | Yes | Yes | Yes | No |
| Yang et al. (2025) | China | NR | 160 | 6 months and 12 months | Nurse-led lifestyle intervention including cardiac rehab and lifestyle modification, emotional counselling, medication adherence, and monitoring of symptoms | Yes | No | No | Yes (SF36) |

*AF: atrial fibrillation. AFSS: AF Severity Scale. ASTA: Arrhythmia-Specific Questionnaire in Tachycardia and Arrhythmia. BP: blood pressure.*
*BMI: body mass index CV: cardiovascular. DCCV: direct current cardioversion. DOAC: direct oral anticoagulant. MI : myocardial infarction.*
*NR: not reported. QOL: quality of life. SF 36: short form 36 questionnaire TIA, transient ischaemic attack.*

**Table 2.** Baseline patient demographics of included randomised controlled trials.

| Trial<br>(Publication<br>year) | No. of<br>participants<br>(LRFM/UC) | Mean Age<br>(LRFM/UC) | Female<br>Gender in %<br>(LRFM/UC) | Persistent AF<br>(LRFM/UC) | HTN in %<br>(LRFM/UC) | DM in %<br>(LRFM/UC) | CHF in %<br>(LRFM/UC) | CAD in %<br>(LRFM/UC) | Stroke/TIA<br>in %<br>(LRFM/UC) | CHADSVASC<br>≥2 in %<br>(LRFM/UC) | Mean BMI<br>(LRFM/UC) | Baseline LA<br>volume in ml<br>(LRFM/UC) | LVEF<br>(LRFM/UC) |
| --- | --- | --- | --- | --- | --- | --- | --- | --- | --- | --- | --- | --- | --- |
| Hendriks et al<br>(2012) | 356/356 | 66/67 | 44.7/37.9 | 68/44 | 52.5/54.2 | 14/12.9 | 7/7 | 9.3/10.7 | 12.4/12.6 | 35.7/35.4 | 27.1/27.3 | NR | 57±10/56±12 |
| Bowyer et al<br>(2017) | 22/19 | 58.3/63.9 | 23.8/42.1 | 7/5 | 31.8/26.3 | NR | 9.1/5.3 | NR | NR | NR | 28.27 | NR | NR |
| RACE4 (2020) | 671/683 | 64/64 | 33/35 | 166/140 | 49/46 | 11/9 | 14/10 | 6/5 | 8/6 | 58/56 | 28/28 | 65±31/67±30 | 55±9/55±9 |
| NEAT (2020) | 36/36 | 63/66 | 44.4/45.7 | NR | 63.9/74.3 | 16.7/17.1 | 13.9/11.4 | NR | 11.1/14.3 | 72.2/74.3 | 30.3/30.1 | NR | NR |
| Yan et al (2022) | 116/119 | 65/65 | 35.3/42.0 | NR | 62.1/78.2 | 26.7/28.6 | 19.8/18.5 | 41.4/31.9 | 19.8/28.5 | 75/78.2 | 25.7/25.9 | NR | NR |
| Vanharen et al<br>(2023) | 33/32 | 65.9/64.7 | 43.7/45.5 | 10/13 | NR | 2/3 | NR | NR | NR | NR | NR | NR | NR |
| NURSECAT-AF<br>(2024) | 33/33 | NR | NR | NR | NR | NR | NR | NR | NR | NR | NR | NR | NR |
| ARREST-AF<br>(2025) | 62/60 | 62/59 | 31/35 | 38/34 | 71/75 | 12.9/15.0 | NR | 9.7/15.0 | 4.8/10 | NR | 32.8/33.6I | NR | NR |
| Li et al (2025) | 198/194 | 70.9/72.8 | 34.8/28.4 | NR | 76.3/64.4 | 20.7/29.4 | 2.5/4.6 | 44.9/39.2 | 2.0/2.6 | 88.4/91.8 | NR | NR | NR |
| POP-AF (2025) | 75/70 | 62/62 | 33.3/17.1 | 43/42 | 46.7/47.1 | 8.0/8.6 | NR | NR | NR | 53.3/45.7 | 30.1/29.6 | 32±5/34±6<br>(reported as<br>LA volume<br>index) | NR |
| Yang et al (2025) | 80/80 | 65.2/64.5 | 30/32.5 | NR | 60/56.3 | 25/? | 17.5/15 | 22.5/21.3 | NR | 46.3/45 | 26.4/26.1 | NR | NR |

*AF: atrial fibrillation. BMI: body mass index. CAD: coronary artery disease. CHF: congestive heart failure. DM: diabetes melitus. HTN: hypertension. LA: left atrium. LVEF: left ventricular ejection fraction. TIA: transient ischaemic attack.*

Central illustration of study characteristics, methods and main findings.

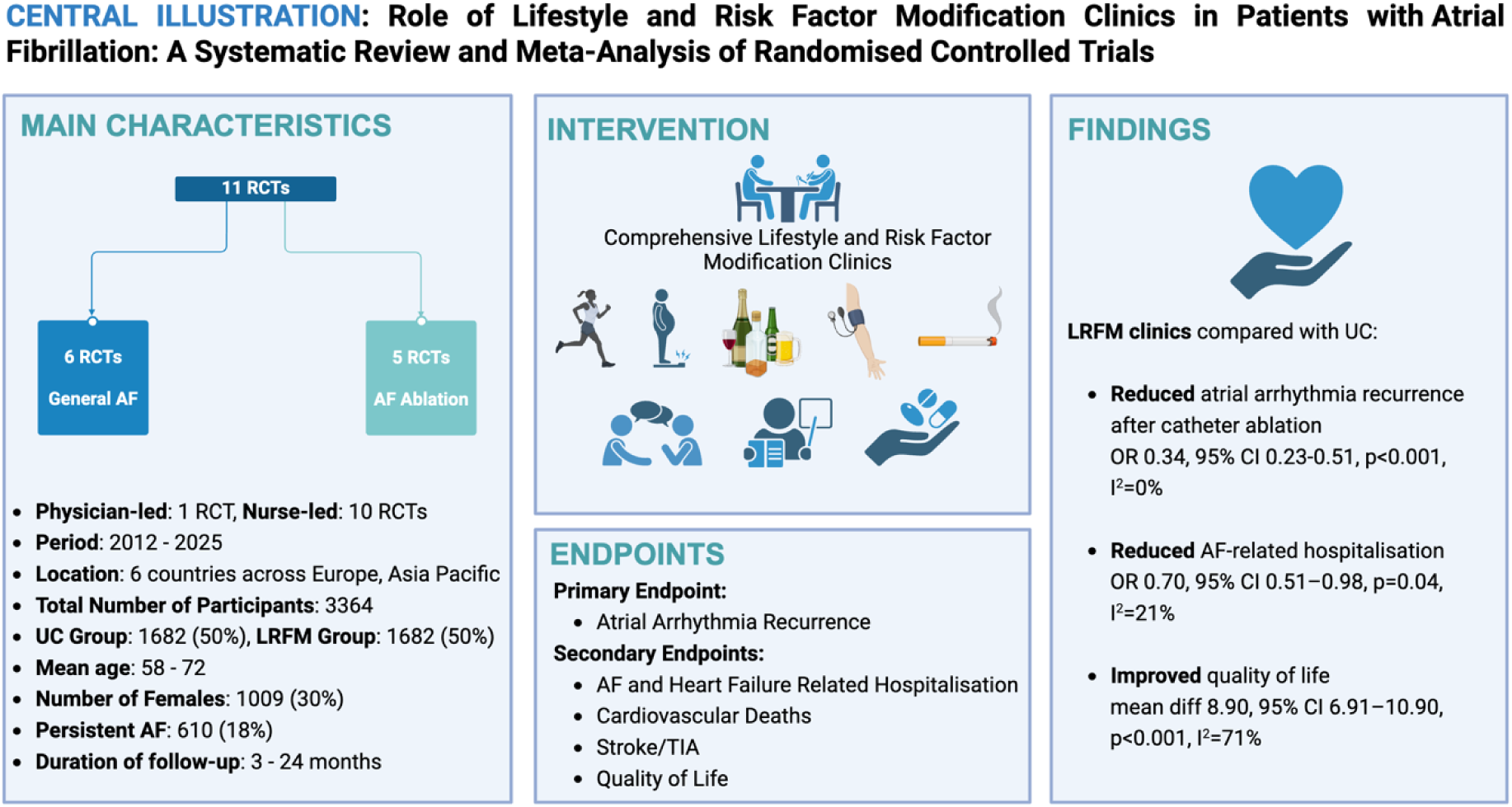

